# Polygenic architecture of rare coding variation across 400,000 exomes

**DOI:** 10.1101/2022.07.06.22277335

**Authors:** Daniel J. Weiner, Ajay Nadig, Karthik A. Jagadeesh, Kushal K. Dey, Benjamin M. Neale, Elise B. Robinson, Konrad J. Karczewski, Luke J. O’Connor

## Abstract

Both common and rare genetic variants influence complex traits and common diseases. Genome-wide association studies have discovered thousands of common-variant associations, and more recently, large-scale exome sequencing studies have identified rare-variant associations in hundreds of genes^1–3^. However, rare-variant genetic architecture is not well characterized, and the relationship between common- and rare-variant architecture is unclear^4^. Here, we quantify the heritability explained by gene-wise burden of rare coding variants and compare the genetic architecture of common and rare variation across 22 common traits and diseases in 400,000 UK Biobank exomes^5^. Rare coding variants (AF = 1e-6 - 1e-3) explain 1.3% (SE = 0.03%) of phenotypic variance on average – much less than common variants – and most burden heritability is explained by ultra-rare loss-of-function variants (AF = 1e-6 - 1e-5). Common and rare variants implicate the same cell types, with similar enrichments, and they have pleiotropic effects on the same pairs of traits, with similar genetic correlations. They partially colocalize at individual genes and loci, but not to the same extent: burden heritability is strongly concentrated in a limited number of significant genes (median: 6 genes explaining 19% of h^2^), while common-variant heritability is much more polygenic. Burden heritability is also more strongly concentrated in constrained genes (median enrichment: 4.5x vs. 2.1x for common variants), indicating that negative selection affects common- and rare-variant architecture differently. Finally, we find that burden heritability for schizophrenia and bipolar disorder^6,7^ is approximately 2%. Our results show that there are a tractable number of large-effect genes to discover by studying rare variants, that common and rare associations are mechanistically convergent, and that rare coding variants will contribute only modestly to missing heritability and population risk stratification.

## Introduction

Genome-wide association studies have discovered thousands of common variants that are associated with common diseases and traits. Common variants have small effect sizes individually, but they combine to explain a large fraction of common disease heritability^8,9^. More recently, sequencing studies have identified hundreds of genes harboring rare coding variants, and these variants can have much larger effect sizes^1–3,5^. However, it is unclear how much heritability rare variants explain in aggregate, or more generally how common- and rare-variant architecture compare: whether they are equally polygenic; whether they implicate the same genes, cell types and genetically correlated risk factors; whether rare variants will contribute meaningfully to population risk stratification.

To characterize common-variant architecture, a productive approach has been to quantify components of heritability by aggregating subtle associations across the genome. This approach has been used to address the problem of “missing heritability”^9–11^, to quantify the shared genetic basis of related diseases and traits^12–14^, to prioritize disease-relevant cell types and regulatory elements^15–18^, and to quantify the effect of negative selection on common-variant architecture^19–22^.

For rare variants, however, heritability estimation is more challenging^23^. Most rare alleles are observed only once or twice, leading to low statistical power, and confounding due to uncorrected population stratification and cryptic relatedness is a major concern. Wainschtein et al.^24^ estimated that common and rare variants combine to explain most of the twin-heritability of height and BMI, with wide confidence intervals, but did not report an estimate for the rare-variant contribution specifically.

To characterize rare variant genetic architecture, we estimated the heritability explained by gene-wise burden of rare and ultra-rare coding alleles, while avoiding confounding due to population stratification. Analyzing association statistics from 394,783 UK Biobank exomes ^5^ together with common-variant association data from the same individuals^25^, we find that the burden heritability due to rare coding variants is modest (1.3% +/-0.03%), and we systematically compare the architecture of common and rare variants.

## Results

### Estimation of burden heritability

In sequencing studies, most rare variants are observed in only one or a few individuals, motivating the use of burden tests that aggregate minor alleles within genes^26^. We define *burden heritability* as the fraction of phenotypic variance explained by minor allele burden in each gene under a random effects model (**Figure 1A**; see Methods). It is a component of the total coding-variant heritability, and it is statistically tractable even for singletons. The alleles comprising the burden are stratified by their predicted functional impact, and we focus primarily on predicted loss-of-function (pLoF) variants, whose gene-wise burden is expected to explain the majority of their total heritability (due to their similar functional consequences) (**Figure 1A**).

**Figure 1:**
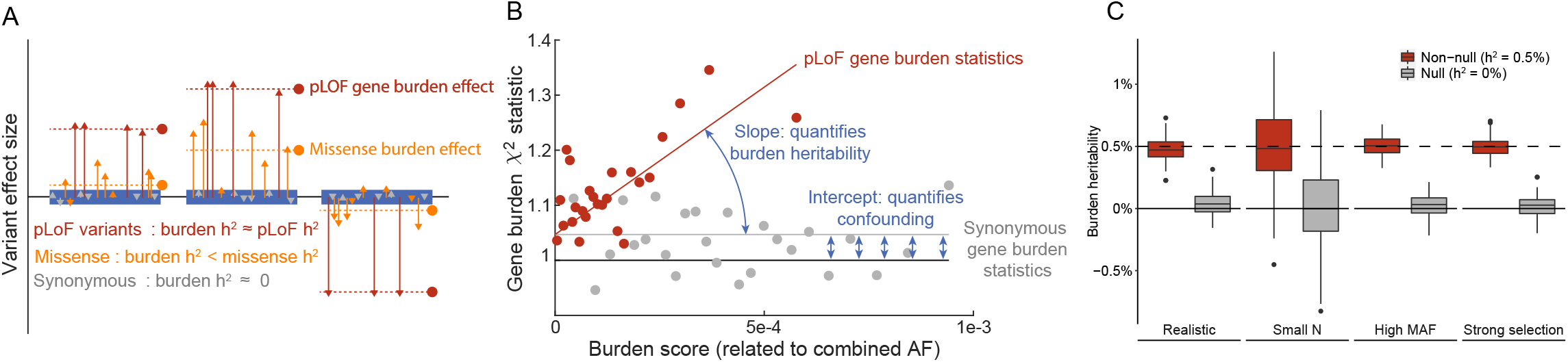
Overview of Burden Heritability Regression (BHR). **(A)** *Burden heritability* is defined as the fraction of phenotypic variance explained by gene-wise burden of minor alleles in a functional class within a certain frequency range (e.g., ultra-rare pLoF variants). The burden heritability of a gene is determined by its mean minor allele effect (dashed lines) and its “burden score,” which is related to the combined allele frequency of those variants. (**B**) BHR regresses gene burden statistics on gene burden scores. The slope of the regression is the mean squared per-allele effect, while the intercept captures confounding factors such as population stratification and sampling noise. We plot real burden scores and effect sizes for ultra-rare pLoF/synonymous variants and LDL, averaging across burden score bins for visualization. (**C**) Performance of BHR in null and non-null simulations. We varied the sample size, the allele frequency of the variants, and the strength of negative selection. The boxplots are the distribution of BHR h^2^ estimates across 100 simulation runs.

We developed *burden heritability regression* (BHR) to estimate burden heritability and to partition it across genes and alleles (see Methods). BHR inputs variant-level association summary statistics and allele frequencies. It regresses burden test statistics on “burden scores,” which are related to the combined allele frequency, and it estimates burden heritability from the regression slope (**Figure 1B**, Supplementary Tables 1-2). Similar to LD score regression^10^, this approach distinguishes heritable signal, which affects the slope of the regression, from confounding due to population stratification and relatedness, which affect its intercept. Intuitively, burden heritability arises from the positively correlated effects of minor alleles within a gene, such that aggregating minor alleles amplifies correlated effects, to an extent that is quantified by the burden score. In contrast, noise and confounding are uncorrelated across alleles, so they are not amplified (however, minor-allele biased stratification is possible; see Methods). BHR relies on the assumption that genes with larger or smaller burden scores do not have larger or smaller per-allele effect sizes, which might be violated due to selection-related effects; we use two approaches to avoid selection-related bias (see Methods). Exome-wide significant genes are excluded from the regression and treated as fixed effects (see Methods). We calculate standard errors using a block jackknife (see Methods). BHR can be used to calculate the heritability and heritability enrichment of subsets of genes by using a different regression slope for each subset (see Methods).

We evaluated the performance of BHR in analyses of simulated data under realistic genetic architectures, with no LD (see Methods, Supplementary Table 3). These simulations included negative selection, which causes trait-associated genes to have fewer minor alleles, and population stratification, which causes systematic inflation in the test statistics. BHR produced unbiased estimates of the burden heritability, and in non-null simulations, it was well powered to detect a burden heritability of 0.5% (**Figure 1C**). We performed an extensive set of simulations to evaluate the robustness of BHR (Supplementary Figure 1), including with different amounts of selection, different amounts of population stratification (including minor-allele biased stratification), different ranges of allele frequencies, and different sample sizes. BHR produced approximately unbiased estimates in all of these simulations.

### Burden heritability of 22 complex traits

We analyzed publicly available UK Biobank exome sequencing association statistics from Genebass for 22 complex traits and up to 394,783 individuals of European ancestry, including 18 continuous traits and 4 common diseases (see Data Availability and Supplementary Table 4) (estimates are reported on observed scale; see Methods)^5^. We analyzed 6.9 million coding variants in 17,318 protein coding genes (see Methods). Within each gene, variants were stratified into three allele frequency bins (MAF < 1e-5, 1e-5 - 1e-4, 1e-4 - 1e-3); we refer to MAF < 1e-5 variants as ultra-rare, and to MAF = 1e-5-1e-3 variants as rare. Variants were also stratified into four functional categories (pLoF, missense damaging, missense benign, and synonymous) (**Figure 2A**, Supplementary Table 5); missense functional predictions were obtained using PolyPhen2^27^

**Figure 2:**
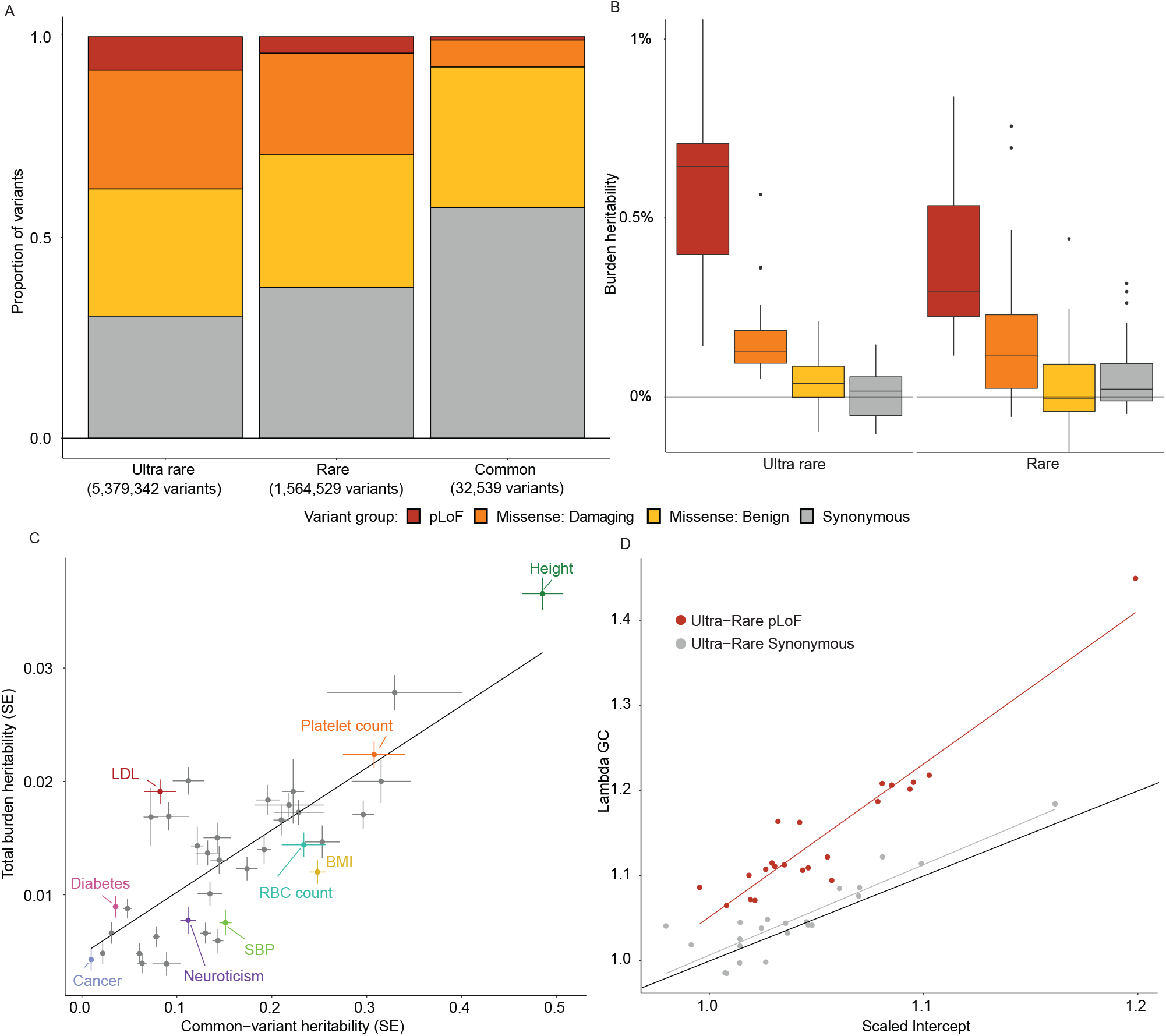
Burden heritability of 22 complex traits and common diseases in UK Biobank. **(A)** Proportions of coding variants by allele frequency and functional consequence in Genebass. Variants in the “Missense: Benign” category are defined by a “benign” Polyphen annotation. Variants in the “Missense: Damaging” category are defined by a “possibly damaging” or “probably damaging” Polyphen annotation. Ultra-rare is defined as AF < 1e-5. Rare is defined as 1e-5 ≤ AF < 1e-3. Common is defined as AF > 0.05 **(B)** Estimates of burden heritability across frequency bins and functional categories. Boxplots show the distribution of heritability estimates across 22 complex traits and common diseases (Supplementary Tables 4, 6). **(C)** Comparison of the total burden heritability (ultra-rare + rare) with the common-variant heritability of each trait (estimated using LDSC^1^). Error bars are standard errors. Numerical results for each trait are contained in Supplementary Tables 7, 8. **(D)** Comparison of test statistic inflation between ultra-rare pLoF (red) and synonymous variants (gray) across the 22 traits. Lambda GC is the median burden χ^2^ statistic divided by 0.454.^2^

We estimate that on average across traits, gene-wise burden of rare and ultra-rare pLoF and damaging missense variants explain 1.3% (SE = 0.03%) of phenotypic variance (**Figure 2B**). All 22 traits had nonzero burden heritability at a nominal significance level (Supplementary Tables 6-7). Burden heritability concentrates among variants with the most severe predicted functional consequences: pLoF variants explain the majority of burden heritability, followed by damaging missense variants, while benign missense variants and synonymous variants explain little or no heritability (**Figure 2B**). Among rare variants, we observed less burden heritability than among ultra-rare variants (median 0.4% across rare pLoF and damaging missense variants). With common-variant summary statistics for the same traits in UK Biobank, we estimated common variant SNP-heritability using LD score regression (see Methods). As expected, a much larger fraction of phenotypic variance is explained by common variants (median 13%), and common-variant and rare burden heritability were highly correlated (**Figure 2C**, Supplementary Table 8).

Inflation in exome association test statistics due to uncorrected population stratification is a major concern, especially when estimating heritability. The BHR intercept quantifies the inflation in burden test statistics due to sampling variation, most forms of confounding, and overdispersion effects (analogous to the LD Score Regression intercept^10^). A potentially problematic form of confounding is minor-allele biased population stratification; however, there was no evidence of this based on the genome-wide average minor allele effect of synonymous variants (Supplementary Figure 2). Across traits, we found that for ultra-rare pLoF variants, confounding and overdispersion explained 4% of the variance in the test statistics, sampling variation explained 85%, and genuine burden heritability explained the remaining 10% (**Figure 2D**, Supplementary Table 6). For ultra-rare synonymous variants, there was zero burden heritability; confounding and overdispersion explained 4% of variance, and sampling variation explained 94% (Supplementary Table 6). These estimates are corrected for within-gene LD, which causes inflation in the burden test statistics in proportion to the number of alleles per gene (see Methods).

### Concentration of burden heritability in significantly associated genes

In GWAS, a consistent observation has been that common diseases and complex traits are highly polygenic: significant loci have small effect sizes, they are spread all across the genome, and they explain a modest fraction of the total common-variant heritability^28,29^. In contrast, most rare diseases are caused by large-effect mutations in a much smaller number of genes, and it is unclear whether the rare-variant genetic architecture of common diseases is highly polygenic like common variants or more oligogenic like rare diseases. We quantified the proportion of burden heritability that is explained by exome-wide significant genes (Methods), and we compared the extent to which common- and rare-variant heritability is concentrated in large-effect genes and regions of the genome.

17 of 22 traits had at least one significantly associated gene in Genebass^5^ (Methods), and they had a median of 6 significant genes per trait (Supplementary Tables 9-10). These genes explained a substantial proportion of the burden heritability (median: 19%; **Figure 3A**), after partially correcting for winner’s curse^30^; see Methods and Supplementary Figure 3. For LDL cholesterol levels, APOB alone explained 39% (SE = 4%) of burden heritability, and for diabetes, GCK explained nearly 15% (SE = 4%). Height had numerous genes explaining >1% of its burden heritability, and even behavioral and cognitive traits had significant genes that explained 1-5%.

**Figure 3:**
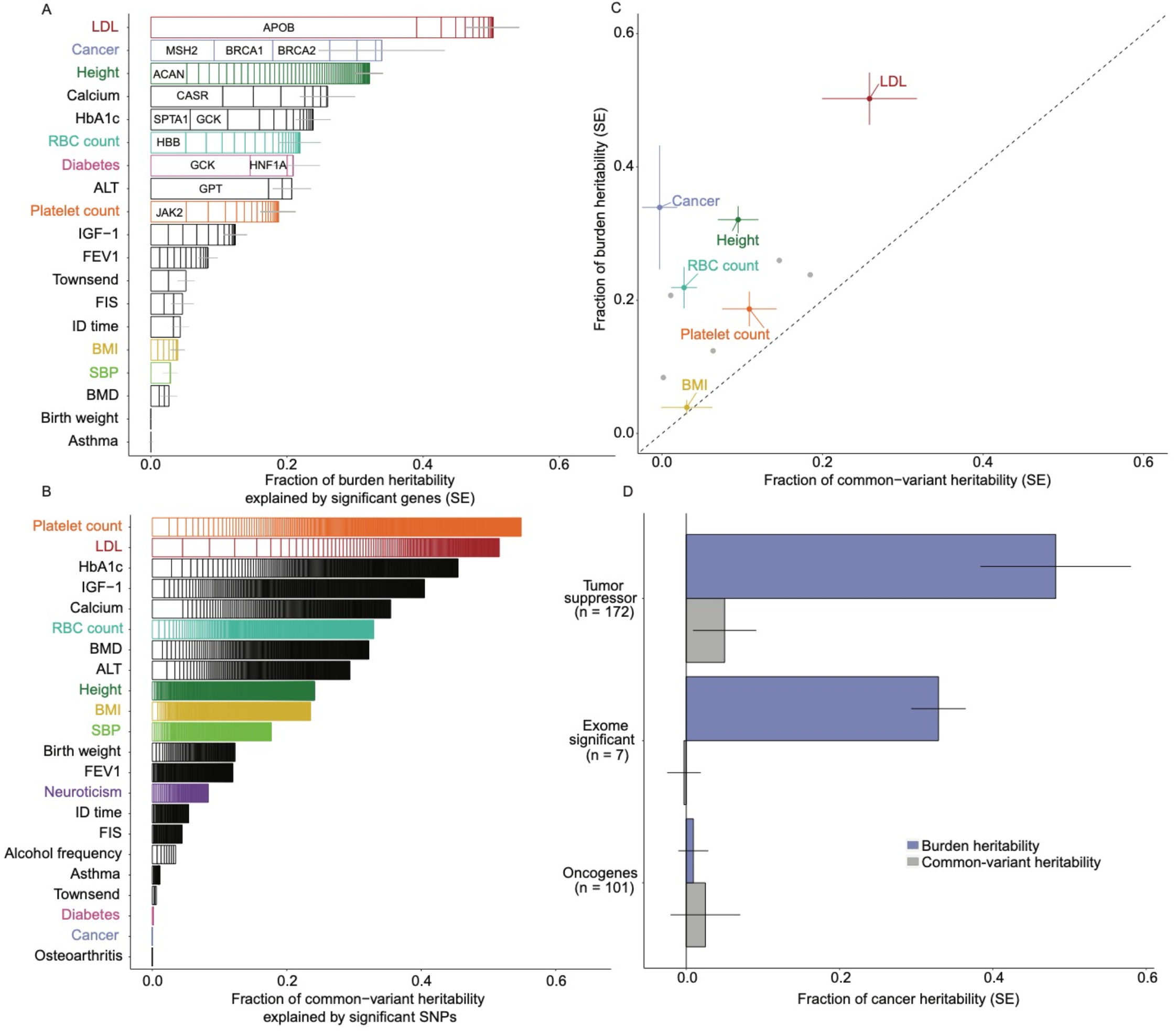
Burden heritability explained by significant genes. **(A)** Fraction of burden heritability explained by exome-wide significant genes from Genebass^3^. Each box represents an exome-wide significant gene for the corresponding trait, and its length represents the fraction of burden heritability it explains. **(B)** Fraction of common variant heritability explained by GWAS significant loci. Each box represents a genome-wide significant locus, and its width represents the fraction of common variant heritability explained by that locus. **(C)** The fraction of common variant heritability mediated by exome-wide significant genes, estimated using AMM^4^, compared with the fraction of burden heritability explained by the same genes, for traits with at least 5 exome-wide significant genes. **(D)** Common-vs. rare-variant cancer heritability mediated by cancer genes. The blue bars are the BHR estimates, and the grey bars are the AMM estimates. Error bars in A-D are standard errors.

In contrast, individual common-variant associations are dramatically smaller as a fraction of common-variant heritability (**Figure 3B**, Supplementary Table 11). Even aggregating common-variant heritability across large LD blocks (most > 1Mb), top rare-variant associated genes (out of 17,318) explain a much larger fraction of heritability than top LD blocks (out of 1,651) (Supplementary Figure 4, Supplementary Table 12). The difference in common-vs. rare-variant polygenicity can be explained by “flattening” due to negative selection, as we previously hypothesized^19^ (see Discussion).

We sought to reconcile the difference in polygenicity with the observation that rare-variant associations are strongly enriched near GWAS loci^3^. For traits with at least 5 significant genes, we quantified the fraction of common-variant heritability mediated by those genes using the Abstract Mediation Model (AMM), which fully accounts for uncertainty in which SNPs regulate which genes^31^ (Supplementary Table 13). We confirm that rare-variant associated genes are enriched for common-variant heritability; for example, the 81 exome-wide significant genes for height explain 9.5% of its common-variant heritability (SE = 2.5%). However, these same genes explain 32.1% of burden heritability (SE = 2.0%) and other traits exhibit similar patterns (median burden:common ratio = 1.9x) (**Figure 3C**). For individual genes as well, LD blocks containing exome-wide significant genes are enriched for common-variant heritability, but these enrichments are modest compared with the rare-variant associations themselves (Supplementary Figure 5). These analyses indicate that while common and rare variants implicate many of the same large-effect genes, rare-variant heritability is more strongly concentrated in those genes, while common-variant heritability is more polygenic.

Five out of the 22 traits (Birth weight, Neuroticism, Alcohol frequency, Asthma, Osteoarthritis; Supplementary Table 9) had significant burden heritability but no individual genes with a significant ultra-rare pLoF burden association. These traits are promising candidates for re-evaluation at larger sample sizes. More generally, none of our estimates for fraction of burden heritability explained by significant genes were close to 1, indicating that there are still substantial opportunities for gene discovery with increased power in sequencing studies.

For the cancer phenotype, which is a composite of multiple cancer types, the seven exome-wide significant genes (MSH2, BRCA1, BRCA2, APC, ATM, PALB2, CHEK2) explain 33% (SE = 4%) of its burden heritability. Noting that all of these genes are well-known tumor suppressors, we analyzed known tumor suppressors and oncogenes from the Cancer Gene Census^32^ (CGC). Indeed, the 172 CGC tumor suppressor genes explain nearly half of the burden heritability (48%, SE = 10%) (**Figure 3D**, Supplementary Table 14). In contrast, the 101 oncogenes do not explain any burden heritability (1%, SE = 2%). These results are concordant with the known biology of tumor suppressors and oncogenes. They contrast with common-variant architecture: tumor suppressor genes only mediate 5% (SE = 4%) of common-variant heritability, and the seven exome-wide significant genes mediate 0% (SE = 2%) (**Figure 3D**).

### Concentration of burden heritability in genes under selective constraint

We investigated the contribution of different gene sets to the burden heritability, defining the burden heritability enrichment of a gene set as its fraction of burden heritability divided by its fraction of burden variance (approximately the fraction of minor alleles, not of genes) (see Methods). We estimated common variant gene-mediated enrichments for the same gene sets using AMM.

First, we analyzed sets of genes that are differentially expressed in trait-matched cell and tissue types (see Methods, Supplementary Tables 10, 14). For these gene sets, burden heritability enrichments and common-variant enrichments were approximately equal (median rare:common enrichment ratio: 1.1x, **Figure 4A**). For example, in a set of 3,396 genes specifically expressed in cordblood-derived red blood cells (RBC), we estimate a 2.1x (SE = 0.4x) enrichment of common variant heritability for RBC count and a 2.2x (SE = 0.3x) enrichment of burden pLoF heritability. This concordance implies that common and rare variation converge on the same causal cell types and tissues. Even though common-variant heritability is less strongly concentrated among top genes compared with burden heritability, it is equally enriched in causal cell types, consistent with the cell-type-centric omnigenic model^33^.

**Figure 4:**
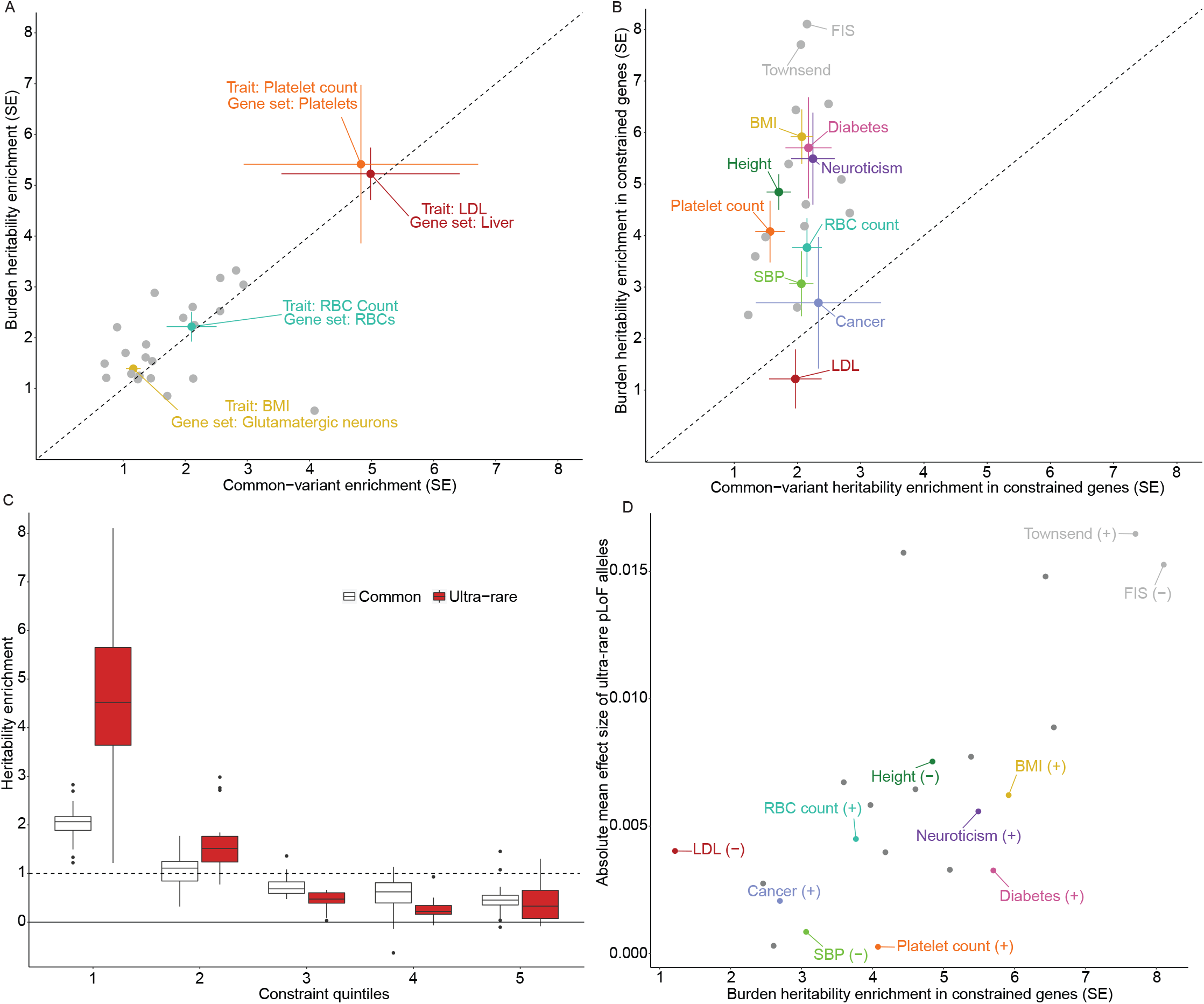
Common-vs. rare-variant heritability enrichments. **(A)** Common and rare variant enrichments across cell type differentially expressed gene sets for selected trait-cell type pairs (see Supplementary Table 14 for the selected trait-gene set pairs). Error bars are standard errors. **(B)** Contrasting common and rare variant enrichments in constrained genes, defined as genes in the bottom 1/5th of observed/expected pLoF alleles in gnomAD^5^. Error bars are standard errors. **(C)** Contrasting common and rare variant enrichments for 22 traits across quintiles of constraint (observed/expected pLoF ratio in GnomAD). **(D)** Absolute mean minor allele effect size of ultra rare pLOF variants genome wide, vs. the constrained gene enrichment of each trait. (+) and (-) denote the sign of the mean minor allele effects.

Next, we compared common-vs. rare-variant enrichments across the spectrum of constraint^34^ (Supplementary Tables 6, 13). Rare variant enrichments were larger than common variant enrichments in constrained genes for 21/22 traits (median rare:common enrichment ratio: 2.5x) (**Figure 4B**). Fluid intelligence score had a rare variant enrichment of 8.1x (SE = 1.0x), compared with a common variant enrichment of 2.2x (SE = 0.3x). We hypothesized that rare variant enrichments would decrease rapidly with decreasing constraint. From the 1st to the 5th quintiles of constraint, rare variant enrichments decayed from a median of 4.5x to 0.3x, while common variant enrichments declined from a lower maximum (2.1x) to a similar minimum (0.5x) (**Figure 4C**). These observations are consistent with the expected effect of negative selection, which prevents both coding and regulatory variants affecting highly constrained genes from becoming common in the population, thereby limiting the magnitude of common variant enrichments^19,22,35,36^ (see Discussion).

For phenotypes that directly affect fitness, loss-of-function alleles are expected to be deleterious almost exclusively, since if gene loss were protective, the gene would be lost. Indeed, pLoF variants in constrained genes are associated with childlessness in UK Biobank^37^. Moreover, a standard approach in severe psychiatric and neurodevelopmental disorders is to aggregate pLoFs across a set of candidate genes^38–40^ (this approach cannot be used to estimate burden heritability, as the candidate-gene burden effect is attenuated when some of the candidate genes are not causal). We calculated the genome-wide mean minor allele effect of ultra rare pLoFs on each trait (Supplementary Table 6). These values were much larger for pLoFs than for synonymous variants (Supplementary Figure 2), indicating that they are not driven by minor-allele biased population stratification (see Methods). Traits with large mean minor-allele effect sizes tended to have a strong burden heritability enrichment in constrained genes (**Figure 4D**), consistent with the hypothesis that these traits are directly under selection (but not providing evidence against the importance of pleiotropic selection^41^).

### Burden genetic correlations across traits and functional categories

Exome-sequencing studies often aggregate pLoF and damaging missense variants to maximize power^6,42^, raising the question of whether damaging missense variants generally act via loss of function. We used BHR to compute burden genetic correlations between pLoF and damaging missense variants (**Figure 5A**, see Methods, Supplementary Table 15). We observed a mean burden genetic correlation of 0.53 (SE = 0.10), implying that pLoF and missense variants in the same genes often have divergent phenotypic effects. One explanation is that deleterious missense variants frequently act via mechanisms other than partial loss of function. Alternatively, PolyPhen2 predictions may vary in quality across genes, such that missense damaging variants approximate pLoFs in some genes but not others.

**Figure 5:**
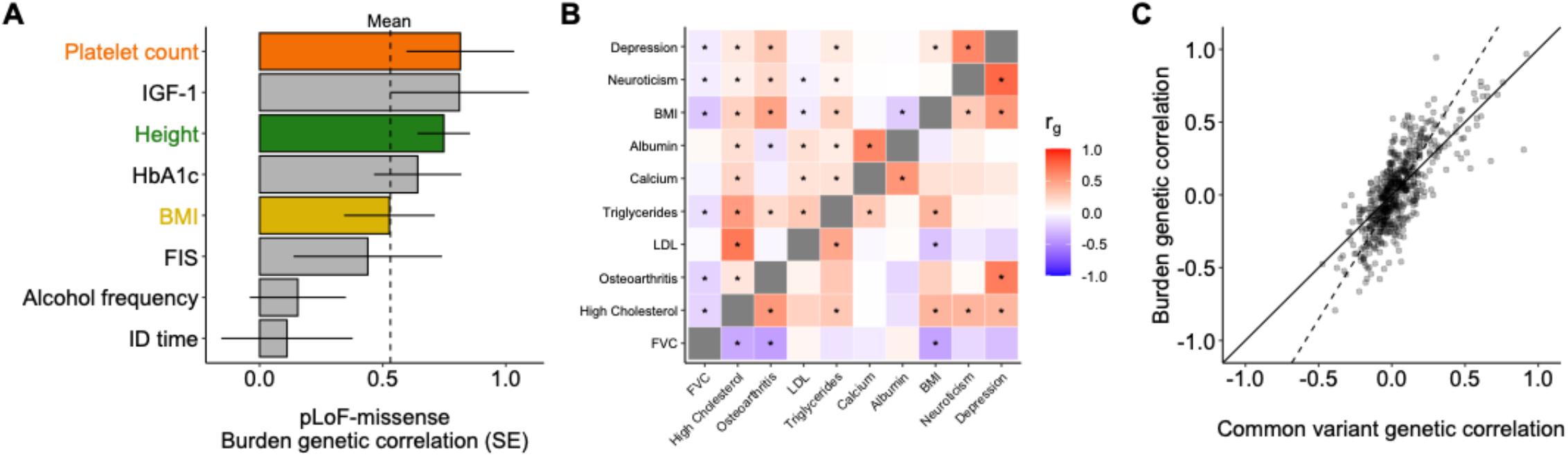
Burden genetic correlations between variant classes and traits. **(A)** Burden genetic correlations between ultra rare pLoF and damaging missense variants, across 9 traits that have nominally significant burden heritability for both classes. Error bars denote standard errors; the mean is 0.53 (s.e.=0.10). **(B)** Clustered heatmap of genetic correlations estimated with Burden Heritability Regression from ultra-rare pLoF variants (lower triangle) and genetic correlations estimated with LD Score Regression (upper triangle). Traits are hierarchically clustered based on BHR genetic correlation. * nominal significance (two-tailed p < 0.05). **(C)** Comparison of common and burden genetic correlations across trait pairs. The dashed line indicates the total least squares regression fit (slope = 1.7).

Common-variant effect sizes are often correlated across traits, providing evidence of shared biological mechanisms. We estimated pairwise burden genetic correlations from ultra-rare pLoF variants among an extended group of 37 traits (Supplementary Table 16). We identified 197 trait pairs that passed a nominal threshold for statistical significance and 55 trait pairs that passed a Bonferroni threshold. For the same group of UKB traits, we also computed common variant genetic correlations using LDSC^12^ (Methods, Supplementary Table 16). We highlight the genetic correlations among 10 selected traits in **Figure 5B**, displaying both burden genetic correlations (lower triangle) and LDSC-computed common variant genetic correlations (upper triangle). Both common and rare variants had correlated effects within clusters of closely related traits (e.g. LDL/Triglycerides/High Cholesterol, Calcium/Albumin, Neuroticism/Depression). and also within less obvious trait pairs (FVC/BMI, Osteoarthritis/Depression).

More generally, rare-variant genetic correlations were concordant with those from common variants (**Figure 5C**). Rare-variant genetic correlations were stronger than common-variant genetic correlations, by 1.7x on average. One potential explanation is that coding variants are less likely to be cell-type specific, and therefore more likely to have pleiotropic effects; a second possibility is that pleiotropic genes are more strongly constrained^41^, which would dampen common-variant genetic correlations. We note that rare-variant genetic correlations, similar to common-variant correlations, can be an artifact of cross-trait assortative mating^43^ (Supplementary Figure 6).

### Burden heritability of schizophrenia and bipolar disorder

Damaging variants in constrained genes are strongly associated with neuropsychiatric disorders^42,44,45^. We applied BHR to estimate neuropsychiatric rare variant genetic architecture using summary statistics from recent exome-sequencing studies of schizophrenia (SCHEMA study^6^: 24,248 cases, 97,322 controls) and bipolar disorder (BipEx study^7^: 14,210 cases, 14,422 controls) (**Methods**). Following the original reports, we analyzed ultra-rare variants with minor allele count less than 5 (MAF < 2e-5 for SCZ, MAF < 9e-5 for BPD).

We estimate that schizophrenia and bipolar disorder have a pLoF burden heritability of 1.7% (SE = 0.3%) and 1.8% (SE = 0.3%) respectively (on a liability scale) (**Figure 6A**, Supplementary Table 17). These estimates were larger than those of the UK Biobank traits except for height, consistent with their high common variant heritability. The burden genetic correlation between bipolar disorder and the two main schizophrenia cohorts was 0.39 (SE = 0.22) and 0.51 (SE = 0.28), roughly consistent with estimates of their common-variant genetic correlation of 0.72^46^. We additionally computed the burden heritability due to ultra-rare damaging missense variants (MPC > 2)^47^, which was significant for schizophrenia (0.35%, SE = 0.12%) but not for bipolar disorder (0.14%, SE = 0.12%). There was no evidence of nonzero burden heritability for synonymous variants.

**Figure 6:**
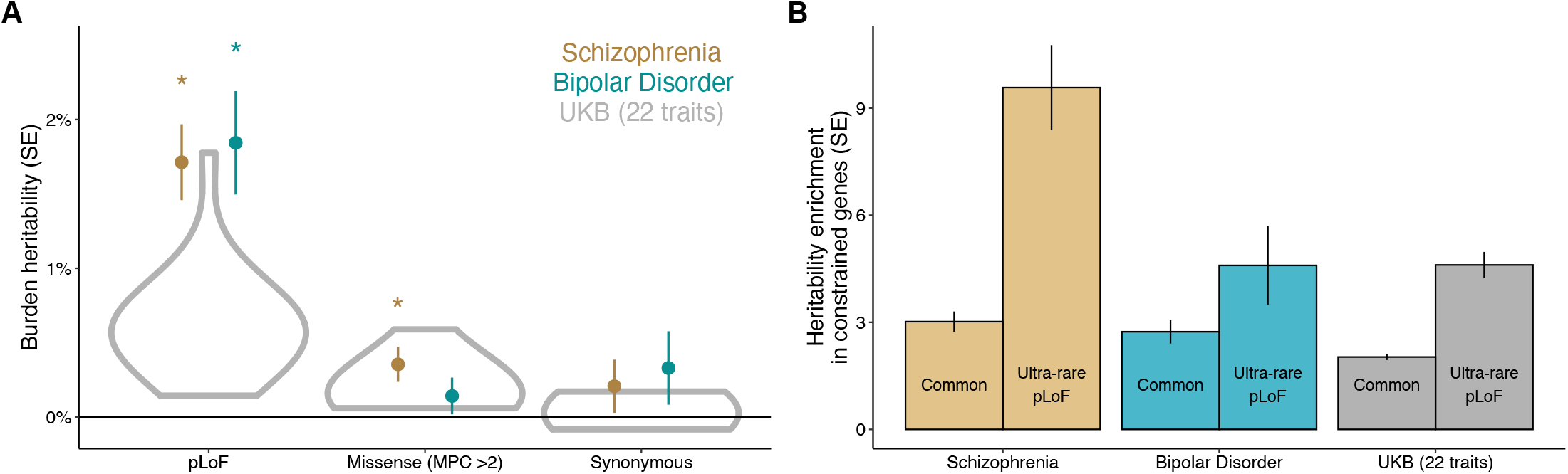
Burden heritability of schizophrenia and bipolar disorder. **(A)** Burden heritability of ultra-rare pLoF variants, ultra-rare missense variants with MPC > 2, and ultra-rare synonymous variants. Gray violin plots show the distribution of burden heritability estimates in 22 UK Biobank traits (Figure 2B). **(B)** Constrained gene enrichment of ultra-rare pLoF vs. common variant heritability. Error bars denote standard errors.

The SCHEMA study^6^ identified 9 autosomal genome-wide significant genes associated with schizophrenia, and we estimate that they explain 7% (SE = 1.5%) of the burden heritability. Larger studies will discover many additional significant genes, and the same will probably occur for bipolar disorder, which has a high burden heritability but no exome-wide significant genes in the BipEx sample.

A consistent observation in exome-based analyses of neuropsychiatric disorders is an enrichment of significant associations in constrained genes^6,7^. Indeed, in the top quintile of constrained genes, burden heritability is 9.6x (SE = 1.2x) enriched for schizophrenia and 4.6x (SE = 1.1x) enriched for bipolar disorder (**Figure 6B)**. The constrained gene enrichment for schizophrenia is the largest observed of any trait in this study; we estimate that constrained genes explain 70% (SE = 9%) of its burden heritability.

## Discussion

Rare protein-coding genetic variation is a rich source of biological insight. Rare diseases are often caused by mutations in one or a handful of genes, and the discovery of those genes has led to effective therapies^48,49^. For common diseases and complex traits, the role of rare variation has been debated^24,50^. In this study, we found that rare loss-of-function variants comprise ∼1% of phenotypic variance for most traits, that burden heritability concentrates among ultra-rare variants in highly constrained, large-effect disease genes, and that these genes are modestly enriched for common-variant heritability. Our findings make us highly optimistic about the potential for rare coding associations to inform our understanding of common disease biology, for two reasons.

First, for rare, syndromic forms of common diseases (e.g., MC4R-driven obesity), a critical question is whether their causal genes are relevant to common variant liability as well. If common and rare variants converge on the same disease-causing processes, therapeutics targeting rare-variant associated genes have the potential to benefit a large number of patients, not only the few who carry specific mutations. Reassuringly, we find that common- and rare-variant associations are mechanistically convergent: rare-variant associated genes are enriched for common-variant heritability (**Figure 3C**), common and rare variants implicate the same cell types and tissues (**Figure 4A**), and they have pleiotropic effects on the same pairs of traits (**Figure 5C**). These findings provide quantitative, genome-wide confirmation of previous reports that common and rare variants implicate overlapping genes^3,6,31^ and pathways^40,51^.

Second, rare-variant architecture is much less polygenic. Already, exome-wide significant genes explain a substantial proportion of the total burden heritability for well-powered traits (**Figure 3A**), suggesting that large-effect mechanisms involve a tractable number of genes. In contrast, common-variant polygenicity has been a challenge for translational efforts^4^. Further work is needed to characterize the distribution of rare-variant effect sizes and the sample sizes that will be needed to identify them.

The differences that we observe between common- and rare-variant architecture support the flattening hypothesis, which we previously proposed as an explanation for extreme common-variant polygenicity^19^. Under the flattening hypothesis, a small fraction of genes and regions of the genome have large effect sizes when mutated, and a much larger number of genes and regions have small effect sizes. Even though small-effect variants are more numerous, the difference in effect size is so large that large-effect genes dominate the rare-variant heritability. However, these genes are constrained, limiting their common-variant associations, and common-variant heritability is spread across a much larger mutational target. This hypothesis provides an explanation for the differences we observe between common vs. rare-variant polygencity (**Figure 3A-B**) and between their enrichments in constrained genes (**Figures 4C, 6B**).

Just as negative selection affects the distribution of heritability among genes, it also affects the fraction of heritability in protein-coding versus regulatory regions. Gazal et al.^22^ found that coding variants explain a much larger fraction of heritability for low-frequency variants (∼26%) than for common variants (∼8%), due to negative selection. If this trend continues at even lower frequencies, then rare and ultra-rare noncoding variants would explain little heritability and would not explain “missing heritability.”

Polygenic risk scores derived from common variants may stratify individuals into clinically meaningful groups^52–54^. The growing accessibility of whole exome and genome sequencing raises the question of whether these genetic profiles should expand to include both common and rare variants. On the one hand, large-effect rare variants can be highly relevant to disease risk for individuals, especially when they have been ascertained by phenotype or family history^55^. However, at a population level, our estimates suggest that rare coding variation will only modestly improve the performance of genetic risk scores^56^.

Our analysis has a number of limitations. First, it is limited to coding variants, and we do not quantify the contribution of rare noncoding variants. Second, for missense variants in particular, burden heritability might represent a fraction of the total rare coding heritability, due to overdispersion effects (**Figure 1A**). We stratified missense variants by their PolyPhen predicted effect, but with a more sophisticated approach, it would be possible to capture a larger fraction of the total missense heritability. Third, our analysis is limited to European-ancestry participants in the UK Biobank, which reflects a well-documented bias in human genetics research^57^. Fourth, the UK Biobank is a relatively healthy population cohort^58^, which limits our power to analyze diseases. For the same reason, the UK Biobank sample might be depleted of deleterious genetic variation^37^, potentially causing decreased burden heritability in this population.

These characteristics of biobanks highlight the importance of ascertained case-control cohorts for the study of disease genetic architecture and the need for public sharing of full variant-level summary statistics from these studies. Sharing of GWAS summary statistics has been catalytic, and we advocate for sequencing studies to share variant-level association statistics, including variant frequencies, functional annotations, and per-allele effect sizes, which are sufficient for estimating genetic architecture with BHR.

Just as genome-wide approaches to common variant associations enabled deeper insight into GWAS data, our approach offers powerful new insight into exome-based association studies. We have released open-source software implementing the full suite of BHR analyses (Code Availability), and we hope that the community will use these tools to further characterize genetic architecture across the variant frequency spectrum.

## Data Availability

All data used in this manuscript is publicly available and documented in Supplementary Tables. All results are available in the Supplementary Tables. Neale Lab UKB GWAS summary statistics: http://www.nealelab.is/uk- biobank/

http://www.nealelab.is/uk-biobank/

https://app.genebass.org/

## Additional information

### Data availability

All data used in this manuscript is publicly available and documented in Supplementary Tables. All results are available in the Supplementary Tables. Neale Lab UKB GWAS summary statistics: http://www.nealelab.is/uk-biobank/

### Code availability

BHR is implemented in R, and its source code is publicly available at https://github.com/ajaynadig/bhr. We have also published scripts allowing the results of the manuscript to be reproduced using publicly available data (Data availability).

## Acknowledgements

The authors are grateful for support from National Institute Mental Health (F30MH129009 to Daniel J. Weiner), National Library of Medicine (T15LM007092 to Daniel Weiner), National Institute of General Medical Science (T32GM007753 to Ajay Nadig), Simons Foundation Autism Research Initiative (704413 to Elise Robinson and Luke O’Connor), and the Broad Institute. We are also grateful to Steven Gazal, Daniel King, Alkes Price, Kaitlin Samocha for analytic assistance and helpful comments on this manuscript. We are grateful to Jinjie Duan for identifying an issue in the first draft of our manuscript.

## Author contributions

D.J.W., A.N. and L.J.O. conceived and designed experiments. K.J. and K.K.D. suggested analyses. B.M.N, E.B.R., K.K. and L.J.O supervised the project. D.J.W., A.N. and L.J.O. performed analyses. D.J.W., A.N. and L.J.O. wrote the manuscript.

## Methods

### Definition of burden heritability

Let *X*_*g*_ be the mean-centered genotype matrix for gene *g*, and let *Z*_*g*_ be the standardized genotype matrix, whose columns have zero mean and unit variance. We define the *burden* for gene *g* as the mean-centered minor allele count for each individual:

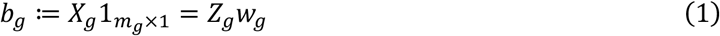

where 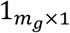 is the all-ones vector, *m*_*g*_ is the number of variants in gene *g*, and *w*_*g*_ is the vector of burden weights. The entries of *w*_*g*_ are the standard deviations of the corresponding columns of *X*_*g*_; under Hardy-Weinberg equilibrium, they are equal to 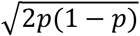 (where *p* is the allele frequency).

Let *y* be the *n* × 1 standardized phenotype vector, and let 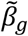 be the *m*_*g*_ × 1 vector of per-allele effect sizes:

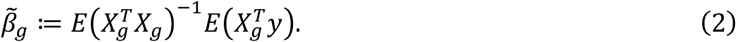

Let *β*_*g*_ be the vector of per-normalized genotype effect sizes, or correlations:

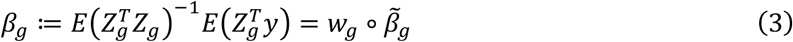

where ∘ denotes the element-wise product. The *burden effect size γ*_*g*_ is the correlation between the burden *b*_*g*_ and the phenotype *y*:

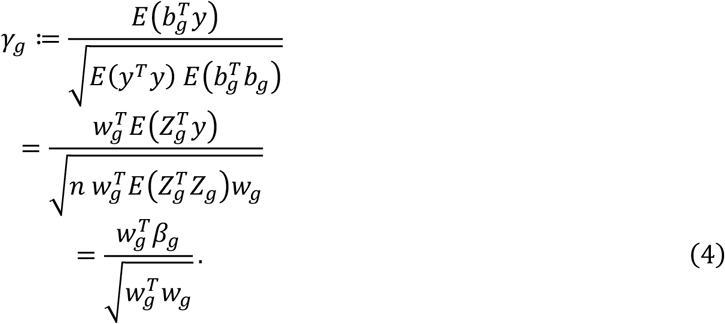

We assumed that there is no LD, such that 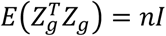, in the third line (see below).

Burden heritability is defined under a random effects model for the burden effect sizes *γ*. Suppose that the vector of per-allele effect sizes 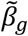 has mean *μ*_*g*_1 and zero covariance. Then the per-normalized genotype effect size vector *β* has mean *μ*_*g*_*w*_*g*_, and the burden effect *γ*_*g*_ has mean

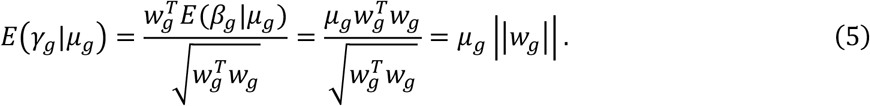

We define the burden heritability of gene *g* as:

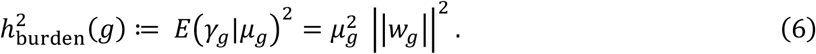

The total burden heritability across a set of genes *A* is:

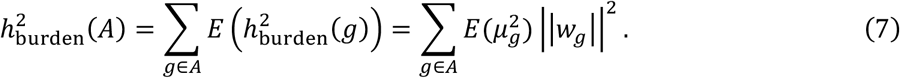

The burden heritability is a component of the total heritability. For gene *g*, its total heritability (without LD) is:

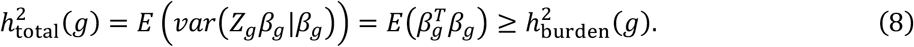

### Burden heritability regression

Burden test statistics, which are commonly used to identify associated genes, are essentially burden effect estimates. The burden effect estimate, *γ*_*g*_, is the sample correlation between *b*_*g*_ and *y*:

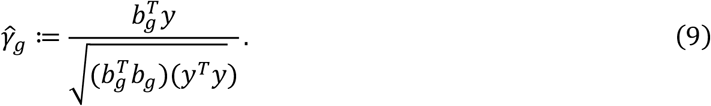

(It is related to the burden χ^2^ statistic: 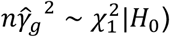. Without LD, and without correlated stratification effects (see below), 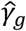 has mean *γ*_*g*_ and variance:

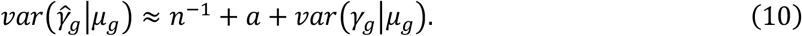

There are three terms. *n*^−1^ is the ordinary sampling variation, which is the approximate term; the approximation is accurate when the burden effect is small. *a* quantifies inflation due to population stratification and cryptic relatedness, and we assume that it is not gene specific (see below). The third term quantifies overdispersion-related sampling variation in the true value of *γ*_*g*_. If variants in the same gene have uncorrelated overdispersion effects with a constant effect-size variance *d*_*g*_ in per-standard deviation units, then the overdispersion term is:

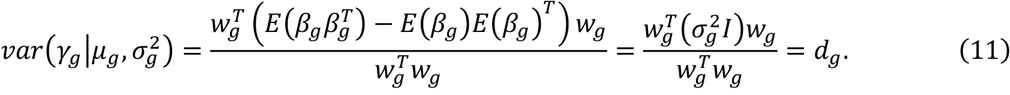

Combining equations 5, 10 and 11:

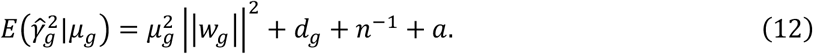

The BHR regression equation is obtained by taking an average value of 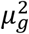 and *d*_*g*_ across genes. Let 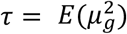 and *d* = *E*(*d*_*g*_). The BHR regression equation is:

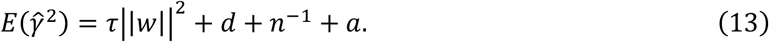

The first term is used to estimate the burden heritability, and the other terms are the regression intercept.

### Minor-allele biased population stratification

A potential source of bias for BHR is minor-allele biased population stratification. Specifically, let *α*_*g*_ be the random vector of normalized stratification effects for minor alleles in gene *g*; we generally assume that

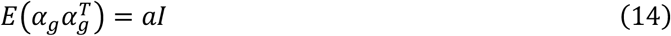

where *a* is the non-gene-specific inflation parameter. Under this assumption, the contribution of stratification to the BHR equation is:

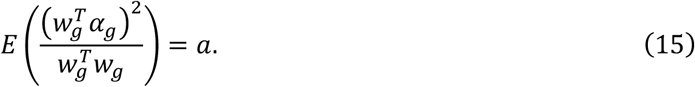

However, minor alleles may have nonzero mean effect sizes due to stratification, such that

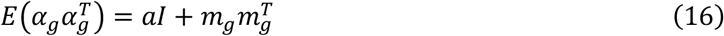

where *m*_*g*_ = *mw*_*g*_ is the mean effect due to stratification. This type of stratification could plausibly arise when a small fraction of individuals in the study come from a certain subpopulation, such that variants specific to that subpopulation are observed at low frequencies. It could also occur when one subpopulation is bottlenecked, causing its frequency spectrum to shift. In this scenario, the contribution of stratification effects is:

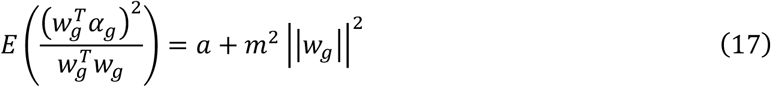

and the BHR slope will be inflated by *m*^2^.

Minor-allele biased stratification would cause the mean minor-allele effect size to be nonzero genome wide, possibly motivating a genome-wide mean centering approach. For pLoF variants, however, it is biologically plausible that their causal effect sizes have nonzero mean (especially for traits such as autism and schizophrenia^38–40^. To distinguish between these possibilities, we calculate the genome-wide mean minor allele effect size for pLoF and synonymous variants separately. Let *w*_*genome*_(*syn*) be the concatenated vector of synonymous burden weights across the genome; we compute the genome-wide mean synonymous minor allele effect, 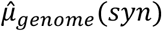, as:

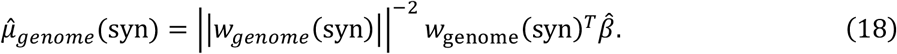

We compare this estimate with the corresponding value for pLoF variants; minor allele-biased stratification is expected to produce nonzero 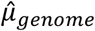 for both, while minor allele-biased causal effects are expected to produce nonzero 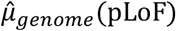 only. In the summary statistics we analyzed, there was no evidence of minor allele-biased stratification (see Supplementary Figure 2). If minor allele-biased stratification is detected, it can be corrected by subtracting 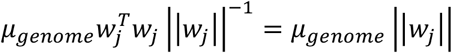 from 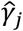.

### Independence assumption and selection-related bias

BHR assumes that 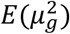 is not correlated with |∣*W*_*g*_ ∣|^2^.In general in a regression analysis, if the slope depends on the independent variable, it leads to bias. Here, the most plausible reason for non-independence is that genes under selective constraint have smaller burden scores and larger mean effect sizes; this would produce downward bias in the heritability estimates.

We use two approaches to mitigate this potential bias. First, we bin genes by their observed vs. expected number of pLoF variants in gnomAD (a measure of selective constraint) [Karczewski et al.]. With this approach, we only require the weaker assumption that 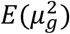 is uncorrelated with |∣*W*_*g*_ ∣|^2^ within bins. We use five bins of approximately equal size. This approach is analogous to the use of LD-related annotations by Gazal et al. to address bias due to LD-dependent architecture in stratified LD score regression (S-LDSC)^21^.

Second, we incorporate null burden statistics that effectively fix the BHR intercept and ameliorate bias in its slope. (Even in the absence of bias, this approach is useful to increase power). We define random null burden weights vectors *ν*_*g*_, whose burden weights are randomly sign flipped compared with *w*_*g*_ (but identical in magnitude). Burden statistics computed using null burden weights are equally affected by noise, confounding, and overdispersion effects, but they contain very little burden signal.

In detail, let *ν*_*g*_ = *w*_*g*_ ∘ [±1, …, ±1] be the null burden weights for gene *g*. The null burden effect size is:

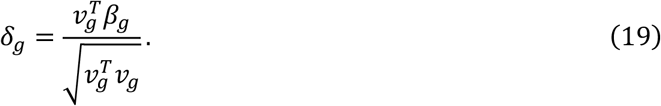

If the mean minor-allele effect is *μ*_*g*_, the mean of *δ*_*g*_ is:

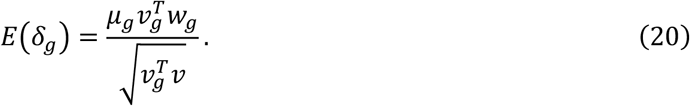

The regression equation for the null burden statistics is

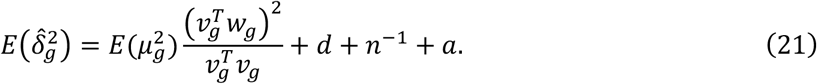

The null burden scores, 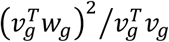, are much smaller than the original burden scores, as the random sign flipping causes 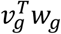 to be small; therefore, the intercept of the regression is effectively constrained to be approximately equal to the mean null burden statistic.

Any number of these null burden statistics can be incorporated into the regression. We use five null burden statistics per gene, which is enough that including a larger number has little effect (Supplementary Figure 8).

### Large-effect genes as fixed effects

Large-effect genes introduce noise in BHR. We identified genes with a significant association at a Bonferroni-significant exome-wide significance threshold (i.e., 0.05 / number of genes by a χ^2^ test). We excluded these genes from the regression and instead included them as fixed effects, adding their squared burden effect size estimates to the heritability directly. This approach is appropriate because the effect size estimates of significant genes are less likely to reflect confounding, and it greatly reduces the standard error of the regression estimator. The estimated heritability explained by each gene was 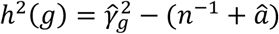, where (*n*^−1^ + *â*) is the BHR intercept.

### Standard errors calculation

We estimated standard errors in the regression using a block jackknife, as previously described^10^. We used 100 contiguous blocks of genes with around 170 genes per block. Significant genes are excluded from the block jackknife procedure, and uncertainty in their effect size estimates is incorporated using the delta method. The delta method is also used to calculate the standard error for the the fraction of burden heritability mediated by significant genes, the enrichment of burden heritability in particular annotations, and the genetic correlation.

In detail, let *θ* be a vector of parameters with covariance matrix Σ. For a function *g*(*θ*), the sampling variance is approximately:

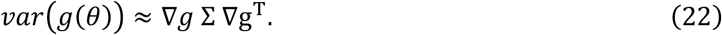

We apply this formula as follows:

- *Standard error of the fraction of burden heritability in a particular gene set*. Let 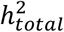 be the total burden heritability estimated by BHR, and 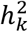 be the burden heritability in annotation *k* estimated by BHR. The fraction of burden heritability in annotation *k* is:

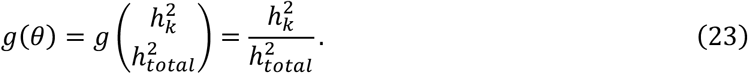

The covariance matrix of *θ*, Σ, is computed via block jackknife.

- *Standard error of the fraction of burden heritability in a particular gene annotation under the mixed model*. In the mixed effects model, genes with exome-wide significant associations are modelled as fixed effects. Let 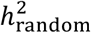 be the total burden heritability estimated by the BHR random effects model excluding significant genes, and let 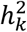 be the burden heritability in annotation *k* estimated by BHR, excluding significant genes. Let 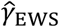 denote the vector of burden effect sizes for exome-wide significant genes. Let *M*(*k*) be the diagonal matrix with dimension equal to the number of significant genes whose diagonal entries are 1 for genes in annotation *k*, and 0 otherwise. The fraction of burden heritability in annotation c is:

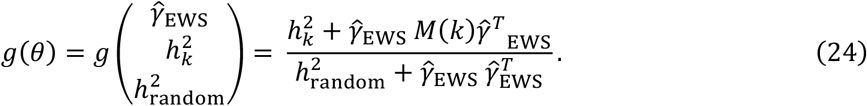

The variances and covariance of 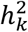 and 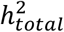 are computed via a block jackknife. The variance of 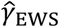 is estimated as the intercept of the BHR random effects model, and their covariance is assumed to be zero with each other and with 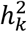 and 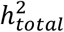.

- *Standard error of the burden genetic correlation under the mixed model*. Let 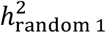 and 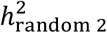 be the random-effects burden heritability of traits 1 and 2 respectively, excluding significant genes. Let *ρ* be the burden genetic covariance between trait 1 and trait 2 excluding significant genes, computed with the cross-trait BHR model. Let 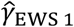 denote the vector of burden effect sizes for significant genes (in per-s.d. units) for trait 1. Let 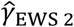 denote the vector of burden effect sizes for significant genes for trait 2. The burden genetic correlation under the mixed model is:

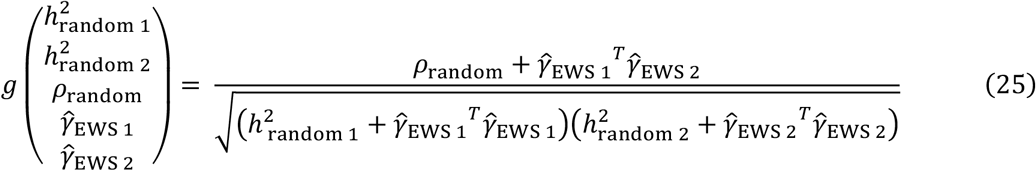

The variances and covariances of 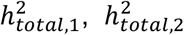, and *ρ* are computed via a block jackknife. The estimated variances of 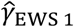 and 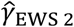 are the BHR intercepts for traits 1 and 2 respectively, and the covariance between 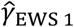 and 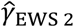 is the BHR cross-trait intercept. The exome-wide significant effects are assumed to have no covariance with 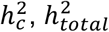, or *ρ*. The covariance between *u* and *ν* is the intercept from the cross-trait BHR model.

### Stratified regression equation and heritability enrichment

BHR can be used to model any number of gene-level annotations. Let *A*_*g*_ be the row vector of annotation values for gene *g*. Similar to S-LDSC^16^, we model the effect-size variance of gene *g* as a linear function of *A*_*g*_:

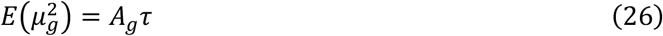

where *τ* is the regression slope. This choice is necessary in order for *τ* to be estimated using linear regression (other choices give rise to least-squares estimators without closed-form solutions). The gene-stratified regression equation becomes

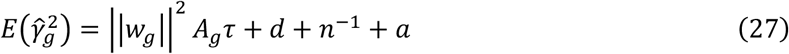

where we assume that overdispersion and confounding effects do not vary across gene sets.

We define the burden heritability enrichment of a gene set as the fraction of heritability divided by the fraction of burden scores. Let the cumulative burden score for gene set *k* be

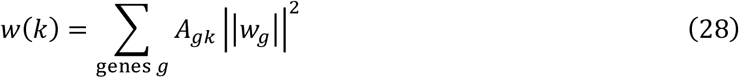

and let its estimated burden heritability be

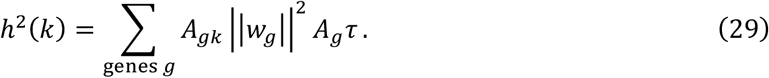

Letting *w*(0), *h*^2^ (0) denote the cumulative burden score and the burden heritability across all genes, the burden heritability enrichment of gene set *k* is:

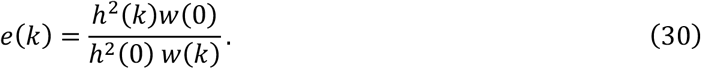

This definition differs from the fraction of heritability divided by the fraction of genes; for example, constrained genes have smaller burden scores on average, so their burden heritability enrichment is greater than their fraction of heritability divided by their fraction of genes.

### Simulations

We simulated gene burden statistics under realistic genetic architectures without LD. We simulated 18,000 genes with between 1 and 1,000 possible variants per gene (drawn from a uniform distribution). We chose the mean effect size for each gene, *μ*_*g*_, from a sparse mixture of normal distributions. In simulations with overdispersion, we also included nonzero gene-specific effect-size variance parameters, *d*_*g*_. Then, we drew per-allele effect sizes for variants within each gene from gene-specific normal distributions:

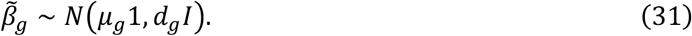

To model negative selection, we simulated effect sizes on 100 independent traits, and we defined a selection coefficient for each variant in proportion to its sum of squared effect sizes across traits. This choice follows the stabilizing pleiotropic selection model of Simone et al.^41^ The selection coefficients were scaled to a desired mean selection coefficient.

We sampled allele frequencies from the neutral spectrum, such that the probability of observing an allele at allele count *n*_*j*_ was proportional to 1/*n*_*j*_, where 1 ≤ *n*_*j*_ ≤ *n*. We approximated the effect of selection of the allele frequency spectrum by discarding variants whose sampled allele frequency was 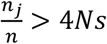, where *s* was the selection coefficient and *N* was 1e4. This approach allows millions of variants to be sampled efficiently.

After sampling the allele frequencies *p*, we set the per-normalized-genotype effect sizes to 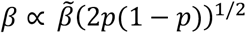, normalizing them so that the burden heritability of variants in the allele frequency bin under consideration matched the desired value.

We computed the observed over expected number of variants in each gene by dividing the number of variants with frequency greater than zero by the number of variants (between 1 and 1000), computing o/e bins from these values.

We sampled effect-size estimates for each variant from a normal distribution, which is appropriate for a continuous trait:

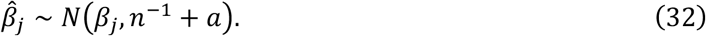

Simulation parameters for each simulation are provided in Supplementary Figure legends 1. For the “realistic” simulations, the fraction of causal genes with large, medium and small effects was 4e-4, 2e-3, 1e-2 respectively, and their per-allele effect size variance before normalization was 5, 1, 1/5. The mean selection coefficient was 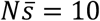. The sample size was 5e5, the number of genes was 1.8e4, and the true burden heritability was either 0 or 0.005. The variance of the stratification effects was 1e-7, the mean minor-allele-biased stratification effect was 1e-5, and there was no overdispersion.

### Observed-scale effect sizes for binary traits

For binary traits, we used raw allele counts in cases in controls as the input to BHR, rather than effect-size estimates from a mixed model. We calculated the observed-scale effect size of SNP *x* on the phenotype *y* as:

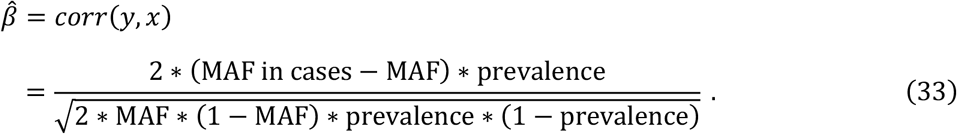

We report heritability estimates on an observed scale unless noted otherwise.

### Genes analyzed

We analyzed 17,318 genes, a subset of the 19,407 genes in Genebass^5^. We analyzed genes meeting all of the following criteria: autosomal; LoF observed/expected ratio present in gnomAD^34^; cell type specific t-statistic defined in Finuncane 2018 Nature Genetics; and at least one variant present in Genebass.

### Variant functional annotations

We analyzed variants in four functional categories: predicted loss-of-function (pLoF), missense pathogenic, missense benign, and synonymous variants. pLoF variants were defined as in Genebass^5^, and included stop-gained, essential splice and frameshift variants. Missense functional classes were defined using PolyPhen2: we defined missense pathogenic as a PolyPhen variant annotation of “probably damaging” or “possibly damaging” and missense benign as a PolyPhen variant annotation of “benign”. Synonymous variants were defined as in Genebass^5^

### Common-variant heritability estimates

We used GWAS summary statistics from the UK Biobank to facilitate a direct comparison with the phenotypes from exome-sequencing analysis (see URLs). Across 22 core BHR traits, the GWAS had a median effective sample size of 344104 (see Supplementary Table 8).

We used stratified LD Score Regression (S-LDSC)^10,16^ to generate common variant heritability and genetic correlation estimates. We elected to use LDSC for direct comparison of heritability estimates because it employs a similar random-effects model to BHR. We used LD scores from the 1000 Genomes project ^10^ and annotations from the baseline LD model^21^ (see URLs).

In order to estimate the fraction of common-variant heritability explained by significant genes (Figure 3), we used HESS, which is able to estimate the local heritability explained by regions with significant associations or significant genes^29^. We used an LD reference panel from the 1000 Genomes Project^10^ and a genome partition composed of approximately LD-independent blocks from^12^.

We used the Abstract Mediation Model (AMM^31^) to estimate the fraction of heritability mediated by gene sets. In brief, AMM estimates the fraction of heritability mediated by a gene set while accounting for uncertainty in SNP-gene mapping. Instead of relying on SNP-to-gene mapping using expression data like eQTLs, AMM first learns a genome-wide probabilistic SNP-to-gene mapping from the decay in heritability across gene proximity (i.e. 27% of heritability mediated by the closest gene). We applied AMM twice: to estimate the fraction of heritability mediated by BHR-significant genes (Figure 3D) and to estimate the enrichment of heritability mediated by constrained genes and gene sets defined by tissue and cell-type expression data (Figure 4A-C). We used a SNP-to-gene probability distributed learned from constrained genes in Weiner et al^31^, which are well-powered across a range of traits.

### Accounting for LD

Rare variants, and to a lesser extent ultra-rare variants, may have within-gene LD. Within-gene LD is a problem for BHR because it causes sampling errors to be correlated among alleles. In particular, if minor alleles have net-positive within gene LD, then their sampling errors will have net-positive correlations, just as true effects are expected to be correlated. This source of bias is potentially strong, as the sampling variance of the effect sizes is large. Net zero LD, which occurs when correlations are nonzero for particular alleles but zero on average, is less of a problem; it leads to decreased power, but not to bias. Outside-of-gene LD is also only a minor concern, as it is not expected to produce net positive correlations between different minor alleles in the same gene.

Net-positive within gene LD can occur as an ascertainment related artefact of binning on the within-sample allele frequency. Suppose that minor alleles within a gene have a mixture of positive and negative LD, such that their net LD is zero: that is, 1^*T*^*R*1 = 0, where 1 is the all ones vector and *R* is the population correlation matrix. Suppose that we sample *n* haplotypes *X*, compute their within-sample LD matrix 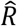, and bin them by their sample minor allele count. For a pair of variants *i,j* with correlation *r*_*j*_, consider the probability that they are both observed exactly *n*_*i*_ = *n*_*j*_ times. This probability is low if *r*_*ij*_ is zero or negative, even if their population allele frequencies are equal. It is much higher, however, if *r*_*ij*_ ≈ 1, which would cause the sample allele frequencies of the two variants to be highly correlated. Conversely, when variants are observed at similar within-sample minor allele frequency, this ascertainment effect makes them more likely to be in positive LD.

If the amount of within-gene LD is known, it can be incorporated into BHR. Let the within-gene-*g* LD matrix be 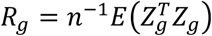. If causal per-allele effect sizes have mean *μ*_*g*_, the mean of the marginal per-normalized-genotype effect size vector *β*_*g*_ is *E*(*β*_*g*_|*μ*_*g*_) = *R*_*g*_*μ*_*g*_*w*_*g*_. The burden effect size is:

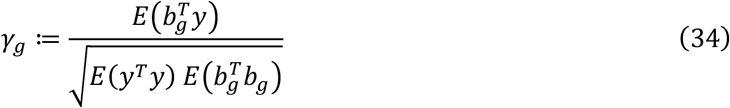

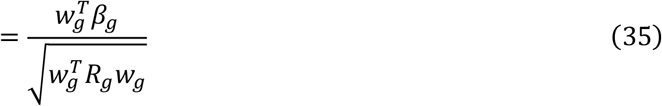

and its mean is:

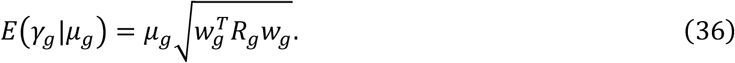

Dropping subscripts, the regression equation becomes:

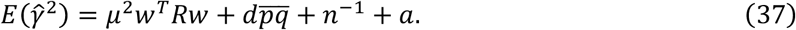

The intercept is unchanged. Let 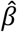 be the vector of sample correlations; their residuals have covariance

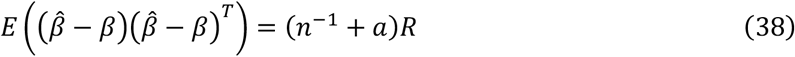

so

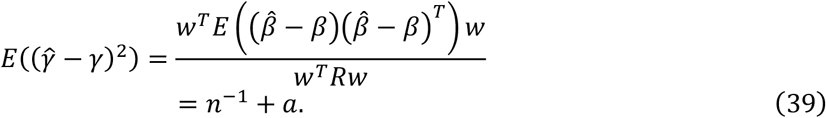

The overdispersion term behaves the same way.

Equation 37 represents one principled approach to account for within-gene LD, but we were only able to access within-gene LD from UK Biobank for chromosomes 20-22. Instead of correcting for within-gene LD using equation 37, therefore, we calculated the amount of bias that is expected to be observed for each class of variants, assuming that the amount of net positive within-gene LD on chromosomes 20-22 are representative of the rest of the genome. Under the null (*β* = 0), the expected burden statistic not accounting for LD is:

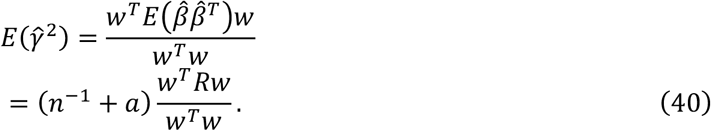

For chromosomes 20-22, we calculated a correction term

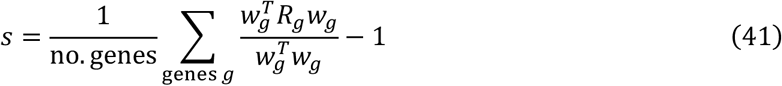

from synonymous variants in each allele frequency bin. The correction factor was noisy for some individual bins and there was no clear relationship between the allele frequency and the correction factor, so we computed a single precision-weighted mean 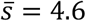 (s.e.=0.5) across bins (see Supplementary Table 20). Then, we subtracted 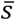 from the BHR regression slope in order to obtain LD corrected heritability estimates; the corrected estimate is equal to

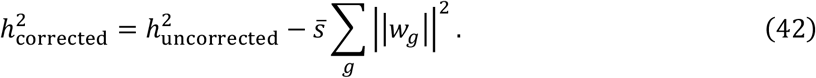

The correction is largest for rare synonymous and missense variants; it is much smaller for ultra-rare variants (which have much smaller entries of *w*) and for pLoF variants (which are fewer in number) (see Supplementary Tables 6, 18). It is inconsequential for ultra-rare pLoFs and in analyses of ultra-rare pLOF variants outside of Figure 2b-c, we do not apply this correction to ultra-rare pLoF estimates.

### Burden heritability explained by exome-wide significant genes

To compute the heritability explained by exome-wide significant genes, we used significant pLoF burden associations (Bonferroni-corrected p < 0.05) from Genebass, which were identified using SAIGE-gene^14^. We computed the ultra-rare pLoF burden statistics for these genes from the SAIGE variant-level effect size estimates (for binary traits, we used the case-control allele frequencies as described above).

When the power to detect a significant gene is smaller than one, its effect size estimate is upwardly biased due to winner’s curse^30^. Similarly, the fraction of heritability explained by significant genes is upwardly biased, especially when most significant genes are close to the significance threshold. We implemented a partial correction for winner’s curse that only depends on the test statistic of each significant gene (and the threshold). In detail, let 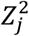 be the χ^2^ test statistic for significant gene *j*, and let *T* be the χ^2^ significance threshold (with 1 degree of freedom). We compute the expected χ^2^ statistic for a gene with non-centrality equal to *Z*_*j*_ conditional on passing the threshold:

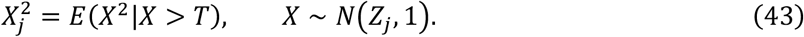

We evaluate the expectation by sampling and computed the winners-curse-corrected test statistic as 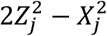.

We tested this approach in simulations and determined that it corrects for about half of the observed winner’s curse across the whole range of genetic architectures and sample sizes (Supplementary Figure 3). It is less successful in the presence of strong population stratification, which causes excess false positives. In real data, a complication is that the significance test is computed from a statistic that includes not only the ultra-rare pLoFs but other variants as well, and this might overcorrection for some genes.

### Genes sets

We analyzed two existing collections of cell type- or tissue-specific gene sets. First, we analyzed tissue-specific gene sets comprising the top 10% of genes differentially expressed in focal tissue vs. other tissues from GTEx v7 bulk RNA-seq^17^. Second, we analyzed cell type-specific gene sets constructed from single-cell RNAseq data^18^. In brief, genes were ranked based on expression in a given cell type relative to expression of the gene in different cell types in the same tissue. Based on the ranking, each gene-cell type pair was assigned a χ^2^ statistic, the statistics were min-max normalized to the range [0,1], and genes with normalized values of 1 were assigned to the gene set.

### Burden genetic correlation

Between two traits, the burden genetic covariance is defined as:

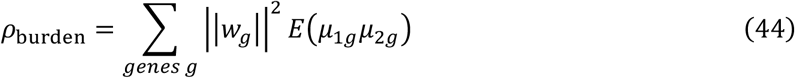

where *μ*_1*g*,_ *μ*_2*g*_ are the mean minor-allele effect size for gene *g* and traits 1 and 2 respectively. The *burden genetic correlation* is:

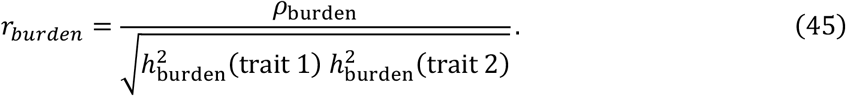

To estimate the burden genetic covariance, the cross-trait BHR regression equation is:

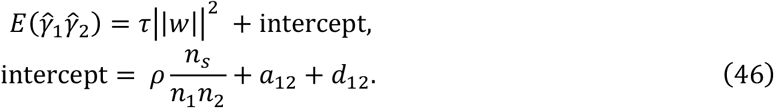

The regression slope is *τ* = *E*(*μ*_1_*μ*_2_), and we stratify the regression across gene sets in the same manner as the single-trait case. In the intercept (similar to cross-trait LDSC^12^), *ρ* is the phenotypic correlation, *n*_*s*_ is the number of samples that are shared between the two studies, *n*_1_ and *n*_2_ are the number of individuals in each study, *a*_12_ is the covariance of the stratification effects on the two traits, and *d*_12_ is the covariance of the overdispersion effects on the two traits.

When the two traits have different sets of variants because there are different individuals for each study, |∣*W*∣|^2^ is replaced by |∣*w*_1_∣| |∣*w*_2_∣| where *w*_*k*_ is the burden weights vector for trait *k*. The same approach is used when computing the correlation between missense and pLOF effects.

With the estimated regression slope *τ*, the estimated genetic covariance is:

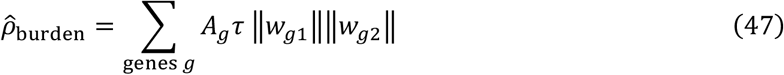

and the genetic correlation is estimated using equation 45.

### SCHEMA and BipEx Datasets

We used publicly available variant-level counts data from the SCHEMA^6^ and BipEx^7^ as input data (URLs). We restricted the SCHEMA analyses to the two study strata with largest sample size: EUR (Exomes, Nextera) and EUR (Exomes, Non-Nextera) (see supplementary information of Singh et al, 2022). For the BipEx dataset, we used the “Bipolar Disorder” group counts. Following Singh et al, 2022, we restricted to variants with minor allele count (MAC) less than 5, and performed separate analyses for pLoF, damaging missense (MPC > 2), and synonymous variants. For each cohort, burden statistics were calculated from allele counts using Equation 33, and burden scores were computed from sample allele frequencies. Then, we used BHR to compute burden heritabilities, enrichments, and genetic correlations separately for the two SCHEMA cohorts. We used this approach to avoid confounding due to differences in the sequencing technology and the sample prevalence between the cohorts.

To produce a single estimate for the schizophrenia heritability, we performed a precision-weighted meta-analysis across the two cohorts. We used BHR to compute the total burden heritability, as well as the burden heritability for constrained genes (the top 5th of genes by observed/expected LoF counts from gnomAD). Within each stratum, we computed the variances for these two estimates, as well as their covariance, using a block jackknife. We used the per-stratum heritability estimates and covariance matrices to perform a precision-weighted meta-analysis. We also computed the jackknife covariance matrix of the heritability estimates for each constraint bin, and used this matrix with the delta method to calculate the standard error for the enrichment of heritability in constrained genes.

## References

1. Sun, B. B. et al. Genetic associations of protein-coding variants in human disease. Nature 603, 95– 102 (2022).

2. Wang, Q. et al. Rare variant contribution to human disease in 281,104 UK Biobank exomes. Nature 597, 527–532 (2021).

3. Backman, J. D. et al. Exome sequencing and analysis of 454,787 UK Biobank participants. Nature 599, 628–634 (2021).

4. Claussnitzer, M. et al. A brief history of human disease genetics. Nature 577, 179–189 (2020).

5. Karczewski, K. J. et al. Systematic single-variant and gene-based association testing of thousands of phenotypes in 426,370 UK Biobank exomes. bioRxiv (2022) doi:10.1101/2021.06.19.21259117.

6. Singh, T. et al. Rare coding variants in ten genes confer substantial risk for schizophrenia. Nature 604, 509–516 (2022).

7. Palmer, D. S. et al. Exome sequencing in bipolar disorder identifies AKAP11 as a risk gene shared with schizophrenia. Nat. Genet. 54, 541–547 (2022).

8. International Schizophrenia Consortium et al. Common polygenic variation contributes to risk of schizophrenia and bipolar disorder. Nature 460, 748–752 (2009).

9. Yang, J. et al. Common SNPs explain a large proportion of the heritability for human height. Nat. Genet. 42, 565–569 (2010).

10. Bulik-Sullivan, B. K. et al. LD Score regression distinguishes confounding from polygenicity in genome-wide association studies. Nat. Genet. 47, 291–295 (2015).

11. Yang, J. et al. Genetic variance estimation with imputed variants finds negligible missing heritability for human height and body mass index. Nat. Genet. 47, 1114–1120 (2015).

12. Bulik-Sullivan, B. et al. An atlas of genetic correlations across human diseases and traits. Nature Genetics vol. 47 1236–1241 (2015).

13. Brainstorm Consortium et al. Analysis of shared heritability in common disorders of the brain. Science 360, (2018).

14. Watanabe, K. et al. A global overview of pleiotropy and genetic architecture in complex traits. Nat. Genet. 51, 1339–1348 (2019).

15. Gusev, A. et al. Partitioning heritability of regulatory and cell-type-specific variants across 11 common diseases. Am. J. Hum. Genet. 95, 535–552 (2014).

16. Finucane, H. K. et al. Partitioning heritability by functional annotation using genome-wide association summary statistics. Nat. Genet. 47, 1228–1235 (2015).

17. Finucane, H. K. et al. Heritability enrichment of specifically expressed genes identifies disease-relevant tissues and cell types. Nat. Genet. 50, 621–629 (2018).

18. Jagadeesh, K. A. et al. Identifying disease-critical cell types and cellular processes across the human body by integration of single-cell profiles and human genetics. bioRxiv (2021) doi:10.1101/2021.03.19.436212.

19. O’Connor, L. J. et al. Extreme Polygenicity of Complex Traits Is Explained by Negative Selection. Am. J. Hum. Genet. 105, 456–476 (2019).

20. Zeng, J. et al. Signatures of negative selection in the genetic architecture of human complex traits. Nat. Genet. 50, 746–753 (2018).

21. Gazal, S. et al. Linkage disequilibrium-dependent architecture of human complex traits shows action of negative selection. Nat. Genet. 49, 1421–1427 (2017).

22. Gazal, S. et al. Functional architecture of low-frequency variants highlights strength of negative selection across coding and non-coding annotations. Nat. Genet. 50, 1600–1607 (2018).

23. Liu, D. J. & Leal, S. M. Estimating genetic effects and quantifying missing heritability explained by identified rare-variant associations. Am. J. Hum. Genet. 91, 585–596 (2012).

24. Wainschtein, P. et al. Assessing the contribution of rare variants to complex trait heritability from whole-genome sequence data. Nat. Genet. 54, 263–273 (2022).

25. Bycroft, C. et al. The UK Biobank resource with deep phenotyping and genomic data. Nature 562, 203–209 (2018).

26. Lee, S., Abecasis, G. R., Boehnke, M. & Lin, X. Rare-variant association analysis: study designs and statistical tests. Am. J. Hum. Genet. 95, 5–23 (2014).

27. Adzhubei, I. A. et al. A method and server for predicting damaging missense mutations. Nat. Methods 7, 248–249 (2010).

28. Loh, P.-R. et al. Contrasting genetic architectures of schizophrenia and other complex diseases using fast variance-components analysis. Nat. Genet. 47, 1385–1392 (2015).

29. Shi, H., Kichaev, G. & Pasaniuc, B. Contrasting the Genetic Architecture of 30 Complex Traits from Summary Association Data. Am. J. Hum. Genet. 99, 139–153 (2016).

30. Palmer, C. & Pe’er, I. Statistical correction of the Winner’s Curse explains replication variability in quantitative trait genome-wide association studies. PLoS Genet. 13, e1006916 (2017).

31. Weiner, D. J., Gazal, S., Robinson, E. B. & O’Connor, L. J. Partitioning gene-mediated disease heritability without eQTLs. Am. J. Hum. Genet. 109, 405–416 (2022).

32. Sondka, Z. et al. The COSMIC Cancer Gene Census: describing genetic dysfunction across all human cancers. Nat. Rev. Cancer 18, 696–705 (2018).

33. Boyle, E. A., Li, Y. I. & Pritchard, J. K. An Expanded View of Complex Traits: From Polygenic to Omnigenic. Cell 169, 1177–1186 (2017).

34. Karczewski, K. J. et al. The mutational constraint spectrum quantified from variation in 141,456 humans. Nature 581, 434–443 (2020).

35. Lek, M. et al. Analysis of protein-coding genetic variation in 60,706 humans. Nature 536, 285–291 (2016).

36. Mostafavi, H., Spence, J. P., Naqvi, S. & Pritchard, J. K. Limited overlap of eQTLs and GWAS hits due to systematic differences in discovery. bioRxiv 2022.05.07.491045 (2022) doi:10.1101/2022.05.07.491045.

37. Gardner, E. J. et al. Reduced reproductive success is associated with selective constraint on human genes. Nature 603, 858–863 (2022).

38. Sebat, J. et al. Strong association of de novo copy number mutations with autism. Science 316, 445–449 (2007).

39. Sanders, S. J. et al. De novo mutations revealed by whole-exome sequencing are strongly associated with autism. Nature 485, 237–241 (2012).

40. Purcell, S. M. et al. A polygenic burden of rare disruptive mutations in schizophrenia. Nature 506, 185–190 (2014).

41. Simons, Y. B., Bullaughey, K., Hudson, R. R. & Sella, G. A population genetic interpretation of GWAS findings for human quantitative traits. PLoS Biol. 16, e2002985 (2018).

42. Fu, J. M. et al. Rare coding variation illuminates the allelic architecture, risk genes, cellular expression patterns, and phenotypic context of autism. bioRxiv (2021) doi:10.1101/2021.12.20.21267194.

43. Border, R. et al. Cross-trait assortative mating is widespread and inflates genetic correlation estimates. bioRxiv 2022.03.21.485215 (2022) doi:10.1101/2022.03.21.485215.

44. Genovese, G. et al. Increased burden of ultra-rare protein-altering variants among 4,877 individuals with schizophrenia. Nat. Neurosci. 19, 1433–1441 (2016).

45. Kosmicki, J. A. et al. Refining the role of de novo protein-truncating variants in neurodevelopmental disorders by using population reference samples. Nat. Genet. 49, 504–510 (2017).

46. Baselmans, B. M. L., Yengo, L., van Rheenen, W. & Wray, N. R. Risk in Relatives, Heritability, SNP-Based Heritability, and Genetic Correlations in Psychiatric Disorders: A Review. Biol. Psychiatry 89, 11–19 (2021).

47. Samocha, K. E. et al. Regional missense constraint improves variant deleteriousness prediction. bioRxiv 148353 (2017) doi:10.1101/148353.

48. Lefebvre, S. et al. Identification and characterization of a spinal muscular atrophy-determining gene. Cell 80, 155–165 (1995).

49. Mendell, J. R. et al. Single-Dose Gene-Replacement Therapy for Spinal Muscular Atrophy. N. Engl. J. Med. 377, 1713–1722 (2017).

50. Pritchard, J. K. Are rare variants responsible for susceptibility to complex diseases? Am. J. Hum. Genet. 69, 124–137 (2001).

51. Kim, S. S. et al. Genes with High Network Connectivity Are Enriched for Disease Heritability. Am. J. Hum. Genet. 104, 896–913 (2019).

52. Khera, A. V. et al. Genome-wide polygenic scores for common diseases identify individuals with risk equivalent to monogenic mutations. Nat. Genet. 50, 1219–1224 (2018).

53. Fahed, A. C. et al. Polygenic background modifies penetrance of monogenic variants for tier 1 genomic conditions. Nat. Commun. 11, 3635 (2020).

54. Khera, A. V. et al. Polygenic Prediction of Weight and Obesity Trajectories from Birth to Adulthood. Cell 177, 587-596.e9 (2019).

55. Bishop, S. L., Thurm, A., Robinson, E. & Sanders, S. J. Prevalence of returnable genetic results based on recognizable phenotypes among children with autism spectrum disorder. bioRxiv (2021) doi:10.1101/2021.05.28.21257736.

56. Biddinger, K. J. et al. Rare and Common Genetic Variation Underlying the Risk of Hypertrophic Cardiomyopathy in a National Biobank. JAMA Cardiol (2022) doi:10.1001/jamacardio.2022.1061.

57. Martin, A. R. et al. Clinical use of current polygenic risk scores may exacerbate health disparities. Nat. Genet. 51, 584–591 (2019).

58. Fry, A. et al. Comparison of Sociodemographic and Health-Related Characteristics of UK Biobank Participants With Those of the General Population. Am. J. Epidemiol. 186, 1026–1034 (2017).

59. 1000 Genomes Project Consortium et al. A global reference for human genetic variation. Nature 526, 68–74 (2015).

60. Berisa, T. & Pickrell, J. K. Approximately independent linkage disequilibrium blocks in human populations. Bioinformatics 32, 283–285 (2016).

61. Zhou, W. et al. Scalable generalized linear mixed model for region-based association tests in large biobanks and cohorts. Nat. Genet. 52, 634–639 (2020).

